# Extended thrombolysis in acute ischemic stroke: A Bayesian meta-analysis and umbrella review

**DOI:** 10.1101/2025.09.22.25336273

**Authors:** Ravi Garg, James M. Brophy

**Affiliations:** Silver Cross Neuroscience Institute, New Lennox, IL 60451 United States of America

**Keywords:** thrombolysis, stroke, umbrella review

## Abstract

**Background:** For acute ischemic stroke, national practice guidelines recommend thrombolysis within a 4.5-hour time window from symptom onset. Randomized trials with advanced neuroimaging have now examined extended thrombolytic time windows, and multiple meta-analyses have provided positive endorsements. However, these previous meta-analyses have not fully exploited the available data, examined quality of life, or reported the probability of clinically meaningful effects. This meta-analysis addresses these potential shortcomings.

**Methods:** We performed a i) systematic literature review up to August 1, 2025, of all randomized controlled trials comparing thrombolysis (alteplase or tenecteplase) with the assistance of advanced neuroimaging to standard, non-thrombolytic therapy in acute ischemic stroke patients with unknown time or beyond 4.5 hours since symptom onset and ii) an umbrella review of previous meta-analyses addressing this issue. Our primary outcome was the mean difference in the utility-weighted modified Rankin Scale (uw-mRS) scores. Secondary outcomes were the absolute risk difference (ARD) for minor disability status and mortality. Bayesian random effects meta-analyses were performed assuming a normal-normal hierarchical model with non-informative priors, which permitted probability calculations for benefit, harm, and regions of practical equivalence (ROPE).

**Results:** Six original randomized trials of extended thrombolysis and 7 meta-analyses were identified but none had considered a uw-mRS score or reported direct probability statements regarding benefit or harm. Our primary uw-mRS outcome showed a ROPE probability of 99% with only a 1% probability of a clinically beneficial response with extended alteplase. There was also a 72% probability that mortality increased by at least 1 life / 100 treated with extended thrombolysis.

**Conclusions:** In contrast with previous publications, this meta-analysis and umbrella review highlights the uncertainty of clinical benefits from extended thrombolysis treatment windows and the need for further high-quality research before this treatment is accepted into routine practice.

## Introduction

Based on randomized controlled trials (RCT), national practice guidelines recommend intravenous thrombolysis with alteplase for eligible acute ischemic stroke (AIS) patients if treatment can be initiated within 4.5 hours of symptom onset.^1^ The mechanism of action of thrombolysis is suspected to be arterial recanalization and subsequent improved brain tissue reperfusion. Given this narrow therapeutic time window for treatment, recent RCTs have explored whether advanced neuroimaging can guide treatment decisions when the time of symptom onset is unknown or extends beyond 4.5 hours from symptom onset.

Since meta-analyses can succinctly summarize trials, enhance statistical precision, and have become the pinnacle of the evidence-based medicine pyramid, it is not surprising that multiple meta-analyses on this topic have already been published. ^2-4^ However, previous meta-analyses have ignored valuable information by dichotomizing outcomes, the difference between statistical and clinical significance, quality of life assessments, and the inferential strength of the evidence. The current meta-analysis addresses these shortcomings, providing additional insights into extended thrombolysis use.

## Methods

### Systematic Review of Randomized Trials and Umbrella Review of Previous Meta-analyses

We performed a systematic search using PubMed, EMBASE, EBSCO and Cochrane databases for phase-3 RCTs comparing thrombolysis (alteplase or tenecteplase) to placebo or standard therapy for participants with unknown time or beyond 4.5 hours since symptom onset; and with the assistance of advanced neuroimaging guidance with penumbral or DWI-FLAIR mismatch; without pre-planned thrombectomy; and published in English before August 1st, 2025 (Supplementary Appendix 1). We excluded observational studies; studies utilizing thrombolytics other than alteplase or tenecteplase; or different imaging techniques. We also systematically searched the databases for eligible meta-analyses (Supplementary Appendix 1). We used Google Scholar to manually review citations from the eligible individual original individual RCTs to identify any additional studies or meta-analyses (Supplementary Appendix 1).

A umbrella review of these meta-analyses was used to confirm the original RCTs and their risk of bias was assessed with the modified Cochrane risk-of-bias instrument.^5^ Publication title, year, sample size, participant characteristics, intervention details, and primary outcome were abstracted for each original individual RCT.^6-11^ We evaluated all the meta-analyses using the critical domains from AMSTAR 2 critical appraisal tool and the Grading of Recommendations Assessment, Development and Evaluation (GRADE) framework.^12,13^

This study followed PRISMA guidance ^14,15^ (SFigure 1). We prepared and published a protocol for this systematic review.^14,15^ The protocol was amended on 5/5/2025 to include tenecteplase and exclude meta-analyses with non-randomized studies.

### Outcome measures

Unlike previous meta-analyses, our primary outcome was the mean difference in utility-weighted modified Rankin Scale (uw-mRS) scores. This represents a quality of life weighted average of the universally reported modified Rankin Scale (mRS) score and reflects patients’ value of their disability status.^16^ Contrary to mRS scores, higher uw-mRS values indicate improved outcomes (uw-mRS 1 equates to mRS = 0 and uw-mRS 0 equates to mRS = 6). We performed sensitivity analyses using different mRS utility weights.^17,18^

Also, unlike most previous meta-analyses, we used the ordinal form of this outcome scale which provides additional insights over the typical dichotomized interpretations of mRS (0-1 versus >1). We established a minimal clinically important difference (MCID) threshold of ≥.09 on the uw-mRS scale based on anchor-based MCID estimations for health utility values among stroke survivors.^19-22^ This MCID represents the smallest possible change in the weighted 7-level scale that reflects a meaningful change between two adjacent health states.

Secondary outcomes were absolute risk differences (ARD) for minor disability status (mRS = 0-1) and mortality (mRS=6). For these secondary outcomes involving the mRS scale, based on recent non-inferiority margins from thrombolytic trials ^23,24^ and national clinical practice guidelines^25^, we considered the MCID to be 3% for a minor disability outcome and 1% for mortality. Based on these MCIDs, regions of practical equivalence (ROPE) were defined as a mean difference of >-.09 to <.09 on the uw-mRS scale; an ARD of >-3% to <3% for minor disability status; and an ARD of >-1% to <1% for mortality.

### Bayesian Meta-analysis

Bayesian analyses allow the formulation of direct probability statements not only for the superiority of treatment over control but also for MCID and ROPE probabilities. We performed a Bayesian random effects meta-analysis for the primary and secondary outcomes assuming a normal-normal hierarchical model. For the primary effect size (the mean difference in uw-mRS scores), we used a non-informative proper normal prior centered at 0 with a standard deviation of 1 (N(0, 1)) thereby allowing the observed data to dominate the posterior probability distribution. For the secondary effect sizes (ARD for minor disability and mortality), we again used a non-informative proper normal prior (N(0,1)). For the heterogeneity parameter, we used a half-normal prior with a scale of.5. We also reported prediction intervals, representing the range in which the effect size for a new, hypothetical study would be expected to fall.

A sensitivity analysis for the primary outcome was performed using a weakly informative prior effect size (N(0, 0.04)) based on the average treatment effect for time-based alteplase treatment. ^26^

### Data Availability

All analyses were completed using R version 4.3.1 (© R Foundation for Statistical Computing, 2023) using the “*bayesmeta*” package which offers direct access to quasi-analytical posterior distributions.^27^. The data and code for all computations can be found in Supplementary Appendix 2.

## Results

Our systematic review of extended thrombolytic RCTs identified five eligible RCTs (SFigure 2). An additional eligible RCT was published after our review dates and included in this study. ^11^ Their summary characteristics and risk of bias assessment were presented in Table 1. All RCTs had a high risk of bias in at least one domain.

**Table 1.**
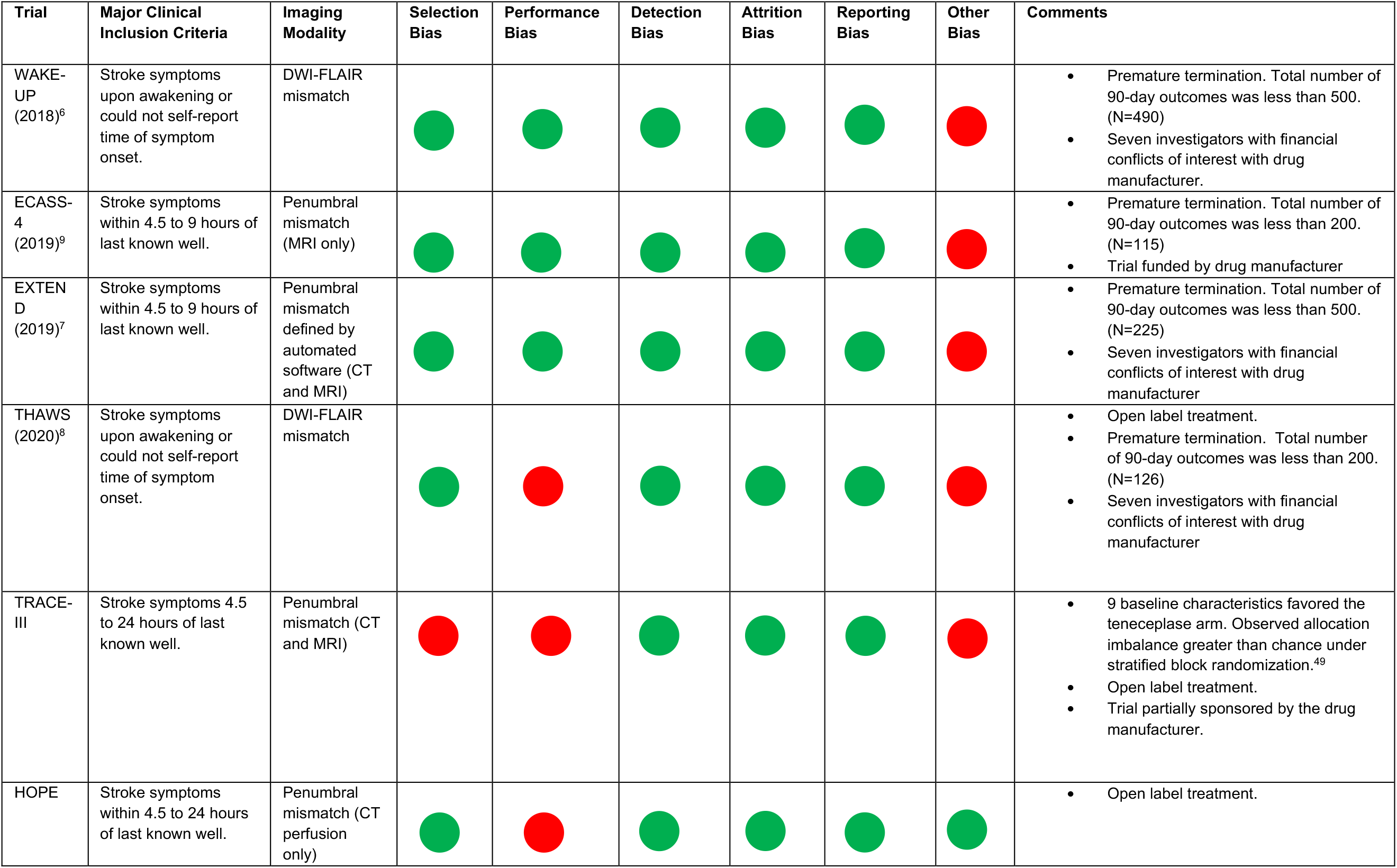
Summary characteristics of recent randomized trials of thrombolysis treatment with neuroimaging guidance. Abbreviations: MRI, magnetic resonance imaging; DWI, diffusion-weighted imaging; FLAIR, fluid-attenuated inversion recovery; hr, hours; CT, computed tomography.

A total of 935 participants were allocated to thrombolysis, and 908 were allocated to control. For each study, we calculated the uw-mRS score, which represents the originally reported mRS outcome values weighted by quality of life measures.^16^ (Table 2).

**Table 2.**
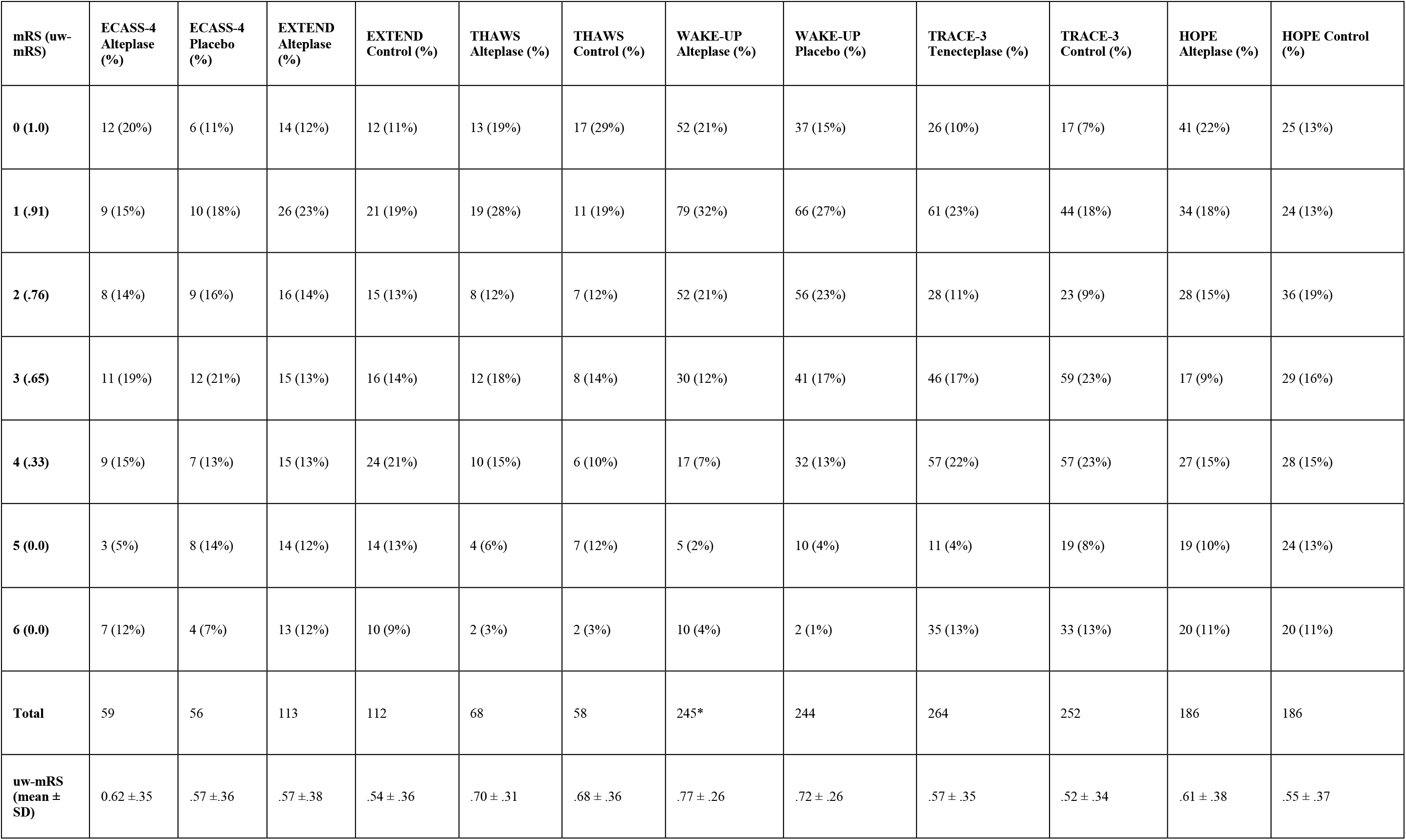
The uw-mRS scores are weighted adjustments of the mRS scores. The calculation of the uw-mRS scores can be found in Supplementary Appendix 2.0. ^*^246 complete cases were reported in the original trial and the discrepancy is due to the estimation from complete cases.

The Bayesian random effects estimate for the primary outcome, the mean difference in uw-mRS, was 0.05 (95% Credible Interval (CrI), 0.01 to 0.08) in favor of thrombolysis (Figure 1). The random effects estimate for the ARD minor disability outcome was.09 (95% CrI,.03 to.15) in favor of thrombolysis; and.02 (95% CrI, -.02 to.05) for mortality in favor of placebo. Forest plots for these secondary outcomes were presented in Supplementary Appendix 3.0. These posterior probability distributions were plotted in Figure 2, with the probabilities of clinically meaningful benefits (green areas), practical equivalence (yellow areas), and harms (red areas) as defined in the methods section, being proportional to the colored area under the curve. The primary outcome of uw-mRS and the secondary outcomes of minor disability showed 98% and 99% probabilities for any benefit, respectively, but only for minor disability was there a high probability of a clinically meaningful benefit (97%). Against this benefit, we noted an 88% probability of increased mortality with thrombolysis with a 72% probability that the mortality increase exceeds 1 life /100 treated. Sensitivity analyses revealed similar results (Supplementary Appendix 4.0). The evidence was summarized in a GRADE evidence profile table (Table 3).

**Table 3.** Grade Evidence Profile. Abbreviations: GRADE = Grading of Recommendations Assessment, Development, and Evaluation; RCT = randomized controlled trial; mRS = modified Rankin Scale.

| *Critical Outcome* | *Risk of bias* | *Inconsistency* | *Indirectness* | *Imprecision* | *Publication bias* | *Probability for clinically significant benefit vs harm* | *Quality of Evidence* |
| --- | --- | --- | --- | --- | --- | --- | --- |
| Critical outcome: Utility-weighted mRS | Serious — all RCTs high risk in $\geq 1$ domain | No serious inconsistency | No serious indirectness | No serious imprecision | Undetected | 1% vs 0% | Low due to risk of bias |
| Secondary outcome: Minor disability (mRS 0–1) | Serious — all RCTs high risk in $\geq 1$ domain | No serious inconsistency | No serious indirectness | No serious imprecision | Undetected | 97% vs <1% | Low due to risk of bias |
| Secondary outcome: Mortality (mRS = 6) | Serious — all RCTs high risk in $\geq 1$ domain | No serious inconsistency | No serious indirectness | No serious imprecision | Undetected | 4% vs 72% | Low due to risk of bias |

**Figure 1.**
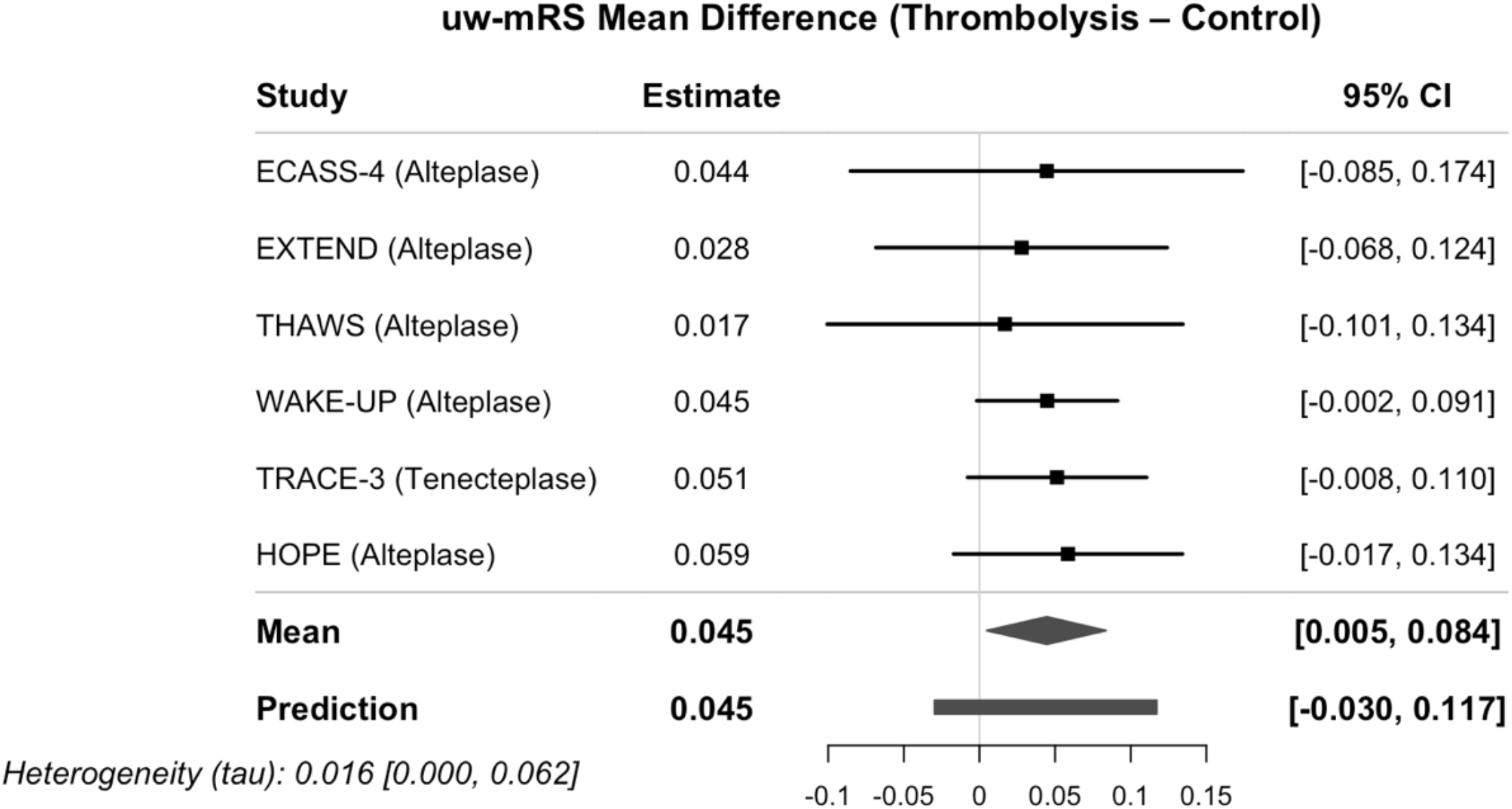
Meta-analysis for mean difference in utility-weighted modified Rankin Scale score.

**Figure 2.**
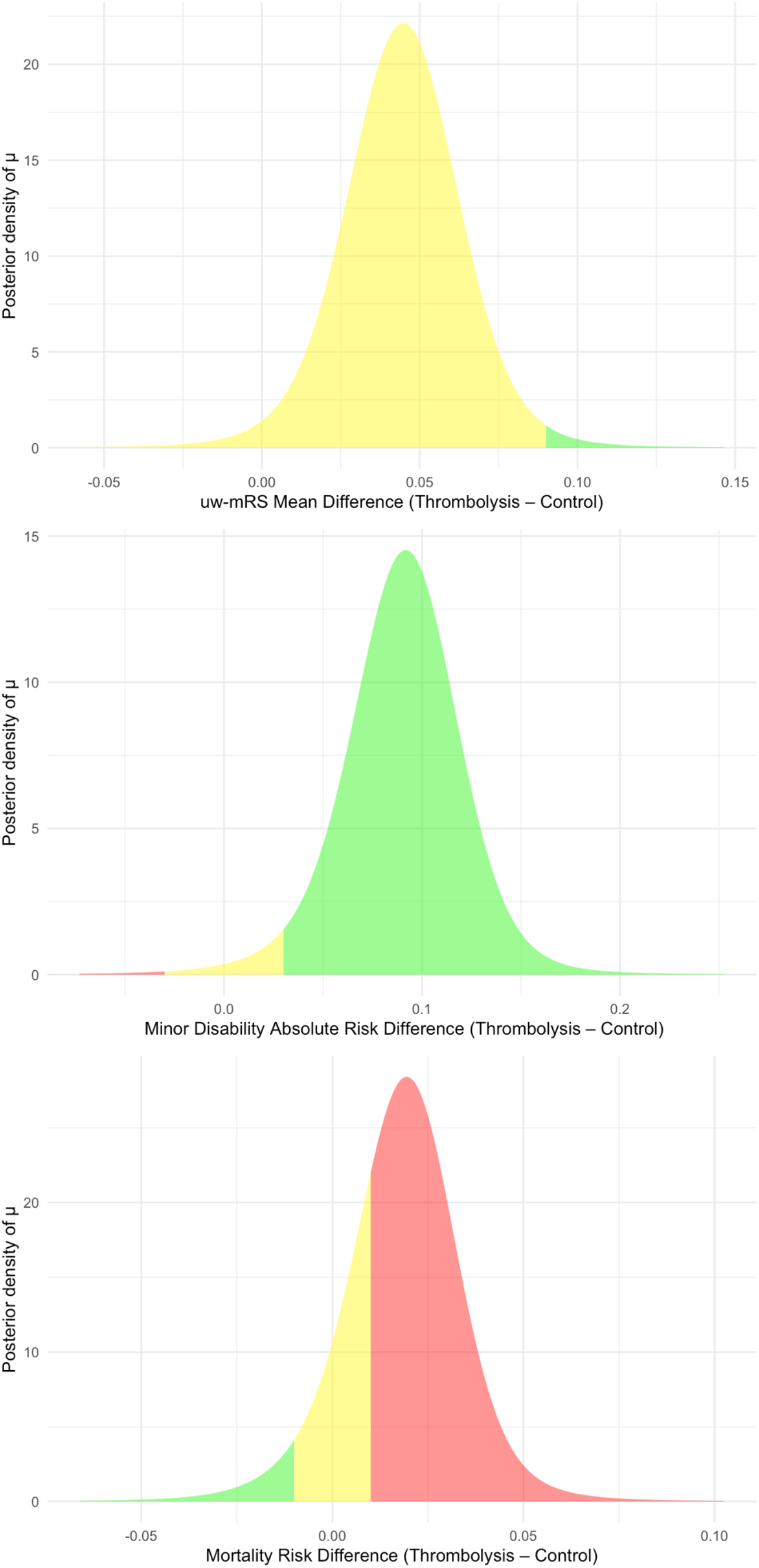
Bayesian Posterior Probability Density Plots. **Figure 2a**. Utility-weighted modified Rankin Scale score outcome. The area under the curve (AUC) in the region of practical equivalence is 99% (yellow area), and clinically significant benefit is 1% (green area). **Figure 2b**. Minor disability outcome. The area under the curve (AUC) in the region of clinically significant harm is <1% (pink area), practical equivalence is 3% (yellow area), and clinically significant benefit is 97% (green area). **Figure 2c**. Mortality outcome. The area under the curve (AUC) in the region of clinically significant harm is 72% (pink area), practical equivalence is 23% (yellow area), and clinically significant benefit is 4% (green area).

A total of 7 previous meta-analyses were identified (SFigure 3). All the meta-analyses used a dichotomized form of the mRS as their critical outcomes^2-4,28-31^ while two meta-analyses did include the ordinal form of the mRS as a secondary outcome.^2,31^ Five of the meta-analyses included phase-2 RCTs with less than 100 participants per group.^3,4,29-31^ Three of the meta-analyses included only subgroups from RCTs in pooled effects.^2,4,31^ Three of the meta-analyses acknowledged early termination as a source of bias (unclear risk).^3,4,31^ One meta-analysis excluded an eligible RCT.^30^ None of the included meta-analyses sufficiently discussed the risk of bias of the included RCTs in their interpretation of the results, defined an MCID or considered financial conflicts of interest (FCOI) as a risk of bias. The full results from the AMSTAR-2 and GRADE critical domains are available in STable 3 and STable 4 respectively.

## Discussion

By examining the totality of the randomized trial evidence, we have shown there is a high probability that extended thrombolysis provides an improvement in the uw-mRS score and minor disability status. However, the probability that this benefit is clinically significant for the uw-mRS outcome is negligible (1%). Against these benefits, we noted a 88% probability of increased mortality and a 72% probability that the mortality is increased by at least one extra death per 100 treated patients

These direct probability statements are possible due to our Bayesian approach and provide more nuanced interpretations of thrombolysis for extended use than reported in previous meta-analyses. Our umbrella review for previous meta-analyses identified 7 publications, which in and by itself is an interesting finding given that only six original RCTs have been performed addressing this question, and all these meta-analyses reached the same favorable conclusion for extended thrombolysis. In addition, these previous meta-analyses had potential sources of systematic error, including an overreliance on subgroup analyses over the entire randomized samples; dichotomization of the ordinal mRS score; and the inclusion of small phase-2 RCTs.

The conventional dichotomization of the mRS score leads to a heterogeneous group of participants who can variously perform complex activities of daily living (mRS=2), basic activities of daily living (mRS=3 and 4), require full assistance with activities of daily living (mRS =5) and participants who died (mRS =6) all being reported under the single outcome rubric of “failures”. ^32^ Conversely, only participants with minimal (mRS =1) or no (mRS =0) disability are deemed “successes”.

These dichotomizations are common in the ischemic stroke literature but are of questionable clinical utility given their wide range of functionality and inconsistency with patient quality of life survey data ^32^. For example, Hanger et al. found a bimodal distribution among a group of stroke survivors and matched controls such that some participants preferred death over any disability while others preferred survival over any disability. ^33^ In another survey, severe hemiplegia was rated worse than death by non-stroke participants.^34^ Conversely, a survey of healthy participants concluded that participants’ preferences varied widely, and few participants viewed the quality of life after a major stroke similar to death. ^35^ We addressed these concerns, at least partially, by adjusting the original ordinal scale results by their respective quality-of-life weights.

Two of meta-analyses included an ordinal analysis of the mRS.^2,31^ Utility-weighted mRS scores differ from an ordinal analysis in several ways. Utility weights reflect different patient valuations assigned to each mRS score compared to an ordinal analysis for which the distance between each score is arbitrary. Under the ordinal assumption, a patient must believe, for example, that transitioning from an mRS score of 6 to 5 has no different value than transitioning from a score of 2 to 1. The effect estimate produced by an ordinal analysis is analogous to, but not numerically equivalent to, an average of the logits for each of the dichotomizations of the mRS. The estimate produced, therefore, may not easily translate to a value that can be used to communicate the risks and benefits of treatment; and has led to concerns about ethical informed consent. ^36^ The limitation in the clinical applicability of the estimate from the proportional odds model has also been described in COVID-19 RCTs employing ordinal outcomes. ^37^ Five previous meta-analysis included phase-2 RCTs with small sample sizes. ^3,29-31,38^ Small sample size (<100 participants per intervention group) has been empirically associated with systematic overestimation of treatment effects of 30-50% in meta-epidemiological research.^39,40^ In all of five meta-analyses, the phase-2 studies had the largest point estimates consistent with this prior research. A further limitation is the use of subgroups in meta-analyses as confounding variables can no longer assumed to be randomly distributed among these smaller subgroups.

Concentrating on subgroup analyses also eliminates the possibility of assessing any treatment effect heterogeneity. For example, in the meta-analysis by Thomalla et al., participants from ECASS-4 and EXTEND were excluded if their time of symptom onset was self-reported.^2^ A more analytically informative way is to assess for treatment effect heterogeneity using the entire sample by symptom onset.

A strength of our Bayesian analysis is its focus on estimation and direct probability inferences. As the complete posterior probability distribution is available, these probability statements are not restricted to specific cut points but can be applied to a range of thresholds, including other personalized MCID choices. This feature is absent from all previous meta-analyses. Our low probability of a clinically significant improvement in overall disability status is consistent with biomarker evidence in RCTs employing MRI for which there was no significant difference in final infarction volume.^6,8^

Other recent RCTs, such as the TWIST^38^ and TIMELESS^41^ RCTs, also support a conservative interpretation of the efficacy of thrombolytics in extended time windows. We excluded these RCTs due to associated clinical heterogeneity from preplanned mechanical thrombectomy.^38,41^ Additionally, the TWIST RCT did not employ advanced neuroimaging. Moreover, neither trial met its primary efficacy endpoint while the point estimate for mortality was higher in the thrombolytic groups consistent with the current findings.

Among the potential limitations of this study, we were obliged to work with aggregate data and although only randomized trials are considered, a chance unbalancing of baseline factors, including baseline disability, may exist due to the relatively small sample sizes. For example, the baseline mRS scores were superior in the alteplase arm of the HOPE RCT; while age and National Institute of Health Stroke Scale (NIHSS) score favored the placebo arm in the EXTEND RCT.

A further limitation is the generalizability of the uw-mRS score weights. However, our conclusions were robust to sensitivity analyses with different utility weights. Finally, Bayesian posterior distributions are a function of the prior distributions and the observed data; and therefore, sensitive to chosen prior distributions. However, our use of non-informative priors ensured that the posterior distributions were dominated by the observed data.

Of course, all estimates obtained from our Bayesian meta-analysis (and all previous meta-analyses) are contingent on unbiased individual trial data in the individual source RCTs. In addition to selection bias and open-label treatment in individual RCTs, we also identified early termination and FCOI as latent sources of bias.

The WAKE-UP RCT, the first RCT prematurely terminated, was terminated not due to efficacy (p=0.02 with nominal alpha = 0.009) but due to loss of funding after randomization of 503 participants of a planned 800 participant enrollment. Consequently, the justification for early termination for THAWS and EXTEND based on WAKE-UP efficacy is questionable.^42^ Three of the seven meta-analyses acknowledged early termination as a potential source of study bias but qualified this risk as unclear.^3,4,31^ However, both simulations and meta-epidemiological studies suggest early termination is associated with a substantial risk of bias. ^43-46^ Comparing completed to truncated RCTs, Bassler et al. found the pooled ratio of relative risks (RRR) was 0.71 (95% CI, 0.65 - 0.77), suggesting truncated RCTs overestimate treatment effects on average by 29% ^43^. These authors also noted the magnitude of over-estimation could increase to 63% (RRR = 0.37 [95% CI, 0.31 - 0.44]) in truncated trials with low events rates (<200 events) as in the present scenario. ^43^ Therefore, based on trial size and, more importantly, event rates, the premature termination of the original RCTs may have resulted in a substantial biased overestimate of the treatment benefit.

Finally, most RCTs had potential FCOI with either direct funding by the drug manufacturer or multiple investigator–manufacturer relationships. Meta-epidemiological data suggest FCOI might systematically bias RCT results with an overestimation of potential benefit.^47^ Naturally, our meta-analysis can’t quantify or adjust for any latent potential biases, but it does suggest that our estimates of benefits, while initially seen as modest, may be overly optimistic.

## Conclusions

In conclusion, our meta-analysis suggests that extended thrombolysis treatment in AIS participants using advanced neuroimaging results has a very low probability of a clinically significant improvement in the uw-mRS score, a high probability of producing a clinically significant improvement in minor disability, and an at least moderate probability of a clinically meaningful increase in mortality. Our umbrella review of previous meta-analyses raises questions about their favorable conclusions. This Bayesian meta-analysis therefore highlights the need for further high-quality research in the form of large RCTs to resolve these substantial uncertainties before this therapy can be employed in routine clinical practice. ^48^

## Data Availability

All data produced in the present study are available upon reasonable request to the authors

## Generative AI declaration statement

The authors did not use a generative artificial intelligence (AI) tool or service to assist with preparation or editing of this work. The authors take full responsibility for the content of this publication.

## Acknowledgements

The authors wish to acknowledge the stroke survivors who participated in these randomized trials.

## CRedIT authorship contribution statement

RG conceived the idea, performed systematic searches, data abstraction, and analysis, and drafted the manuscript. JB assisted with the systematic search, data analysis, and manuscript writing. Both authors contributed to the study design, data interpretation, and critical revision of the manuscript for important intellectual content. RG is the guarantor for the manuscript.

## Ethics

This study did not involve the collection of new human or animal data. The analyses were based exclusively on previously published, de-identified aggregate data from randomized controlled trials and systematic reviews available in the public domain. In accordance with local regulations and institutional requirements, formal ethical approval and informed consent were not required.

