## supplemental file for "Extended thrombolysis in acute ischemic stroke: A Bayesian meta-analysis and umbrella review"

**APPENDIX**

1. Search Strategies
    a. STable1. Search terms used for databases.
    b. SFigure 1. PRISMA 2020 Checklist.
    c. SFigure 2. Systematic search flowchart for the identification of eligible randomized trials.
    d. SFigure 3. Systematic search flowchart for the identification of eligible meta-analyses.
2. Statistical Code
3. Forest Plots
    a. SFigure 4: Bayesian random-effects meta-analysis for a minor disability outcome
    b. SFigure 5: Bayesian random-effects meta-analysis for a mortality outcome
4. Sensitivity Analyses
    a. Posterior effect size applying a weakly informative prior effect size
    b. STable 2: Posterior effect size using different utility weights
5. AMSTAR 2 and GRADE Evaluation
    a. STable 3: AMSTAR 2 Critical Domains
    b. STable 4: GRADE Assessment

**APPENDIX 1: Search strategies**

| Database | Search Terms |
|---|---|
| Pubmed | ((alteplase) OR (tenecteplase)) AND ((stroke)) OR (cerebrovascular accident)) AND ((unknown time of onset) OR (unknown onset) OR (wake-up) OR (extended time window)) |
| Embase | : ('alteplase'/exp OR 'alteplase' OR 'tenecteplase'/exp OR 'tenecteplase') AND ('cerebrovascular accident'/exp OR 'cerebrovascular accident') AND ('unknown time of onset' OR 'unknown time of onset':ti,ab,kw OR 'unknown onset' OR 'unknown onset':ti,ab,kw OR 'wake-up' OR 'wake-up':ti,ab,kw OR 'extended time window' OR 'extended time window':ti,ab,kw) |
| EBSCO | (alteplase OR tenecteplase) AND (stroke or cerebrovascular accident or cva) AND ("unknown time of onset" OR "unknown onset" OR "wake-up" OR "extended time window") |
| COCHRANE | ((alteplase OR tenecteplase) AND (stroke OR cerebrovascular accident) AND (unknown time of onset OR unknown onset OR wake-up OR extended time window)) |

STable1. Search terms used for databases.

| Section and Topic | Item # | Checklist item | Location where item is reported |
|---|---|---|---|
| **TITLE** | | | |
| Title | 1 | Identify the report as a systematic review. | Page 1, Title. |
| **ABSTRACT** | | | |
| Abstract | 2 | See the PRISMA 2020 for Abstracts checklist. | Page 2, Abstract. |
| **INTRODUCTION** | | | |
| Rationale | 3 | Describe the rationale for the review in the context of existing knowledge. | Page 3, Introduction, Paragraph 2. |
| Objectives | 4 | Provide an explicit statement of the objective(s) or question(s) the review addresses. | Page 3, Introduction, Paragraph 3. |
| **METHODS** | | | |
| Eligibility criteria | 5 | Specify the inclusion and exclusion criteria for the review and how studies were grouped for the syntheses. | Page 4, Methods |
| Information sources | 6 | Specify all databases, registers, websites, organisations, reference lists and other sources searched or consulted to identify studies. Specify the date when each source was last searched or consulted. | Page 4, Methods |
| Search strategy | 7 | Present the full search strategies for all databases, registers and websites, including any filters and limits used. | Supplementary Appendix 1. |
| Selection process | 8 | Specify the methods used to decide whether a study met the inclusion criteria of the review, including how many reviewers screened each record and each report retrieved, whether they worked independently, and if applicable, details of automation tools used in the process. | Page 3-4, Methods |
| Data collection process | 9 | Specify the methods used to collect data from reports, including how many reviewers collected data from each report, whether they worked independently, any processes for obtaining or confirming data from study investigators, and if applicable, details of automation tools used in the process. | Page 3-4, Methods |
| Data items | 10a | List and define all outcomes for which data were sought. Specify whether all results that were compatible with each outcome domain in each study | Page 3-4, Methods |

| Section and Topic | Item # | Checklist item | Location where item is reported |
|---|---|---|---|
| | | were sought (e.g. for all measures, time points, analyses), and if not, the methods used to decide which results to collect. | |
| | 10b | List and define all other variables for which data were sought (e.g. participant and intervention characteristics, funding sources). Describe any assumptions made about any missing or unclear information. | Page 3-4, Methods |
| Study risk of bias assessment | 11 | Specify the methods used to assess risk of bias in the included studies, including details of the tool(s) used, how many reviewers assessed each study and whether they worked independently, and if applicable, details of automation tools used in the process. | Page 3-4, Methods |
| Effect measures | 12 | Specify for each outcome the effect measure(s) (e.g. risk ratio, mean difference) used in the synthesis or presentation of results. | Page 3-4, Methods |
| Synthesis methods | 13a | Describe the processes used to decide which studies were eligible for each synthesis (e.g. tabulating the study intervention characteristics and comparing against the planned groups for each synthesis (item #5)). | Page 3-4, Methods |
| | 13b | Describe any methods required to prepare the data for presentation or synthesis, such as handling of missing summary statistics, or data conversions. | Page 3-4, Methods |
| | 13c | Describe any methods used to tabulate or visually display results of individual studies and syntheses. | Page 3-4, Methods |
| | 13d | Describe any methods used to synthesize results and provide a rationale for the choice(s). If meta-analysis was performed, describe the model(s), method(s) to identify the presence and extent of statistical heterogeneity, and software package(s) used. | Page 3-4, Methods |
| | 13e | Describe any methods used to explore possible causes of heterogeneity among study results (e.g. subgroup analysis, meta-regression). | Page 3-4, Methods |
| | 13f | Describe any sensitivity analyses conducted to assess robustness of the synthesized results. | Page 3-4, Methods |
| Reporting bias assessment | 14 | Describe any methods used to assess risk of bias due to missing results in a synthesis (arising from reporting biases). | Page 3-4, Methods |
| Certainty assessment | 15 | Describe any methods used to assess certainty (or confidence) in the body of evidence for an outcome. | Page 3-4, Methods |

| Section and Topic | Item # | Checklist item | Location where item is reported |
|---|---|---|---|
| **RESULTS** | | | |
| Study selection | 16a | Describe the results of the search and selection process, from the number of records identified in the search to the number of studies included in the review, ideally using a flow diagram. | Page 5-13, Results |
| | 16b | Cite studies that might appear to meet the inclusion criteria, but which were excluded, and explain why they were excluded. | Page 5-13, Results |
| Study characteristics | 17 | Cite each included study and present its characteristics. | Page 5-13, Results |
| Risk of bias in studies | 18 | Present assessments of risk of bias for each included study. | Page 5-13, Results |
| Results of individual studies | 19 | For all outcomes, present, for each study: (a) summary statistics for each group (where appropriate) and (b) an effect estimate and its precision (e.g. confidence/credible interval), ideally using structured tables or plots. | Page 5-13, Results |
| Results of syntheses | 20a | For each synthesis, briefly summarise the characteristics and risk of bias among contributing studies. | Page 5-13, Results |
| | 20b | Present results of all statistical syntheses conducted. If meta-analysis was done, present for each the summary estimate and its precision (e.g. confidence/credible interval) and measures of statistical heterogeneity. If comparing groups, describe the direction of the effect. | Page 5-13, Results |
| | 20c | Present results of all investigations of possible causes of heterogeneity among study results. | Page 5-13, Results |
| | 20d | Present results of all sensitivity analyses conducted to assess the robustness of the synthesized results. | Page 5-13, Results |
| Reporting biases | 21 | Present assessments of risk of bias due to missing results (arising from reporting biases) for each synthesis assessed. | Page 5-13, Results |
| Certainty of evidence | 22 | Present assessments of certainty (or confidence) in the body of evidence for each outcome assessed. | Page 5-13, Results |
| **DISCUSSION** | | | |
| Discussion | 23a | Provide a general interpretation of the results in the context of other evidence. | Page 14-16, Discussion |
| | 23b | Discuss any limitations of the evidence included in the review. | Page 14-16, Discussion |
| | 23c | Discuss any limitations of the review processes used. | Page 14-16, Discussion |

| Section and Topic | Item # | Checklist item | Location where item is reported |
|---|---|---|---|
| | | | Page 14-16, Discussion |
| | 23d | Discuss implications of the results for practice, policy, and future research. | Page 14-16, Discussion |
| **OTHER INFORMATION** | | | |
| Registration and protocol | 24a | Provide registration information for the review, including register name and registration number, or state that the review was not registered. | Page 3-4, Methods |
| | 24b | Indicate where the review protocol can be accessed, or state that a protocol was not prepared. | Page 3-4, Methods |
| | 24c | Describe and explain any amendments to information provided at registration or in the protocol. | Page 3-4, Methods |
| Support | 25 | Describe sources of financial or non-financial support for the review, and the role of the funders or sponsors in the review. | Page 18 |
| Competing interests | 26 | Declare any competing interests of review authors. | Page 18 |
| Availability of data, code and other materials | 27 | Report which of the following are publicly available and where they can be found: template data collection forms; data extracted from included studies; data used for all analyses; analytic code; any other materials used in the review. | Supplementary Appendix 2. |

SFigure 1. PRISMA 2020 Checklist.

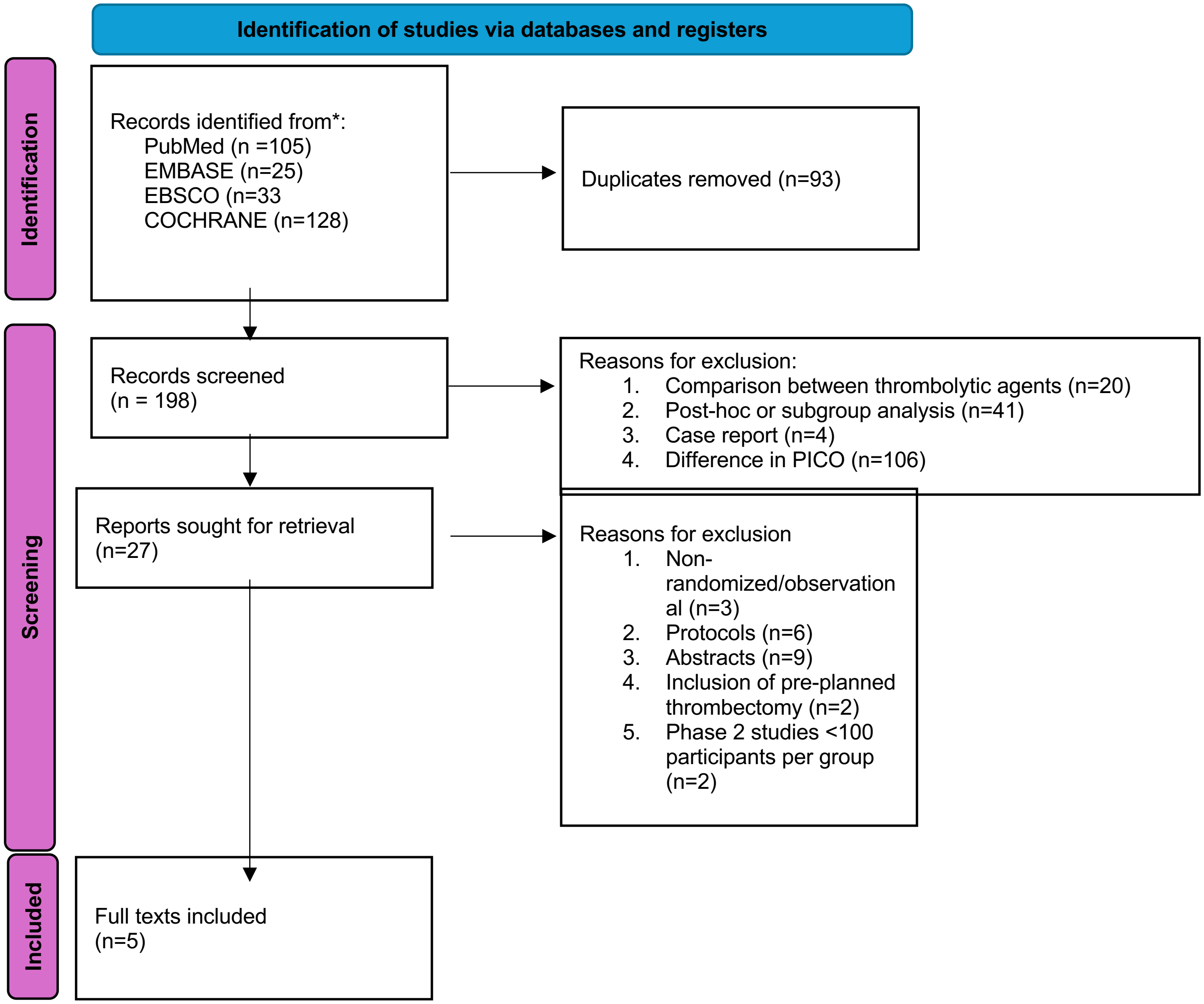


SFigure 2. Systematic search flowchart for the identification of eligible randomized trials.

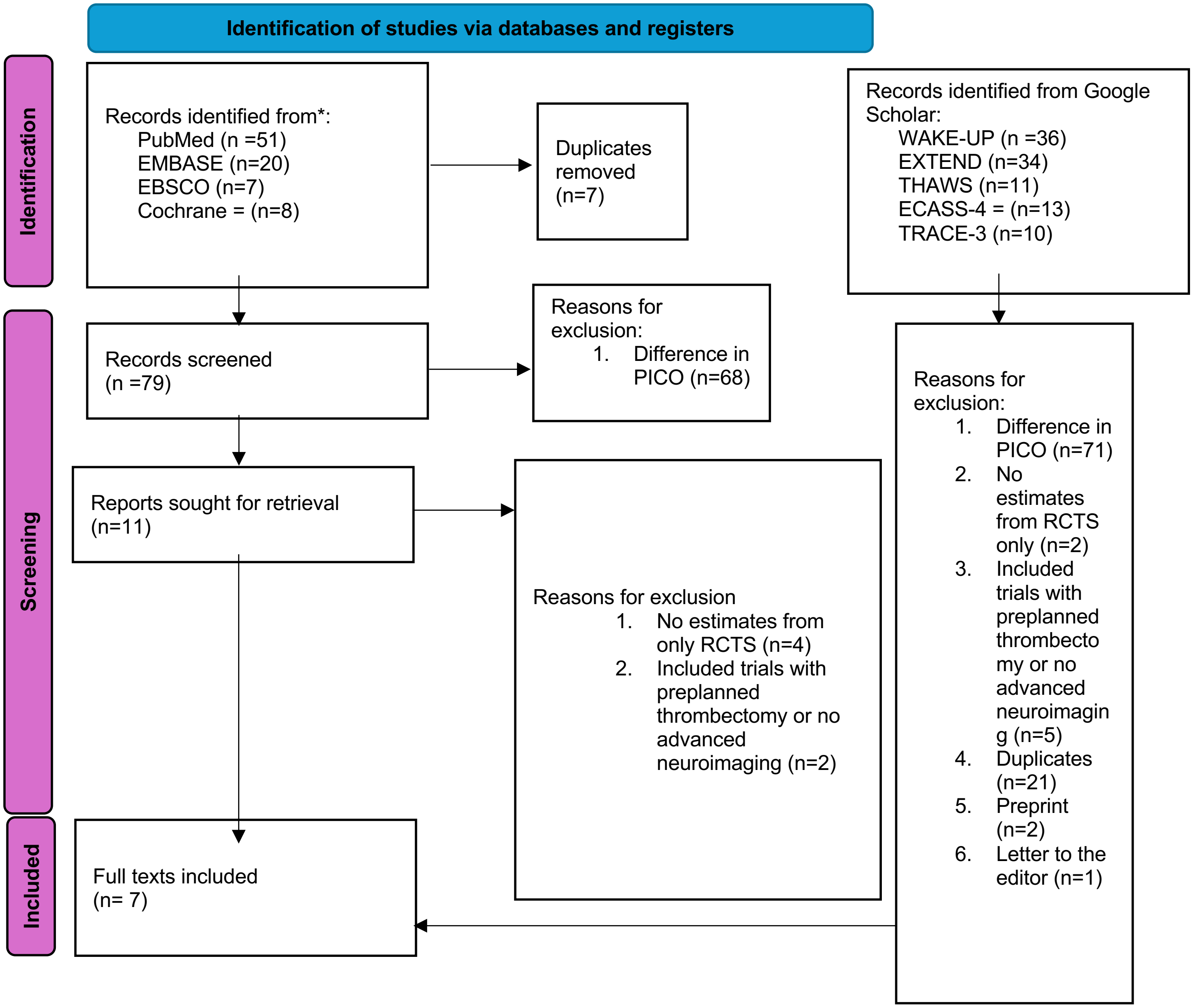


SFigure 3. Systematic search flowchart for the identification of eligible meta-analyses.

**APPENDIX 2: Statistical code**

```
# Libraries
library(bayesmeta)
library(ggplot2)

#  Data from table (counts per mRS category)
# Order within each vector: mRS 0,1,2,3,4,5,6
# Trials: ECASS-4, EXTEND(A), THAWS(A), WAKE-UP(A), TRACE-3 (TNK), HOPE (A)
intervention_counts <- list(
  c(12,  9,  8, 11,  9,  3,  7),  # ECASS-4 Alteplase (n=59)
  c(14, 26, 16, 15, 15, 14, 13),  # EXTEND Alteplase (n=113)
  c(13, 19,  8, 12, 10,  4,  2),  # THAWS Alteplase (n=68)
  c(52, 79, 52, 30, 17,  5, 10),  # WAKE-UP Alteplase (n=245)
  c(26, 61, 28, 46, 57, 11, 35),  # TRACE-3 Tenecteplase (n=264)
  c(41, 34, 28, 17, 27, 19, 20)   # HOPE Alteplase (n=186)
)

control_counts <- list(
  c( 6, 10,  9, 12,  7,  8,  4),  # ECASS-4 Placebo (n=56)
  c(12, 21, 15, 16, 24, 14, 10),  # EXTEND Control (n=112)
  c(17, 11,  7,  8,  6,  7,  2),  # THAWS Control (n=58)
  c(37, 66, 56, 41, 32, 10,  2),  # WAKE-UP Placebo (n=244)
  c(17, 44, 23, 59, 57, 19, 33),  # TRACE-3 Control (n=252)
  c(25, 24, 36, 29, 28, 24, 20)   # HOPE Control (n=186)
)

study_labels <- c(
  "ECASS-4 (Alteplase)",
  "EXTEND (Alteplase)",
  "THAWS (Alteplase)",
  "WAKE-UP (Alteplase)",
  "TRACE-3 (Tenecteplase)",
  "HOPE (Alteplase)"
```

```
)

# Utility weights
weights <- c(1.00, 0.91, 0.76, 0.65, 0.33, 0.00, 0.00)

# Actual posterior (mu|data) shading via bayesmeta$dposterior
plot_posterior_areas_bayesmeta <- function(fit, lower_bound, upper_bound, title,
                                           reverse_colors = FALSE, grid_sd = 5, n = 4001) {
  mu_mean <- fit$summary["mean","mu"]
  mu_sd   <- fit$summary["sd","mu"]
  x <- seq(mu_mean - grid_sd * mu_sd, mu_mean + grid_sd * mu_sd, length.out = n)
  y <- fit$dposterior(mu = x)
  df <- data.frame(x = x, y = y)

  if (reverse_colors) {
    ggplot(df, aes(x, y)) +
      geom_area(data = subset(df, x <= lower_bound), alpha = 0.5, fill = "green") +
      geom_area(data = subset(df, x >  lower_bound & x < upper_bound), alpha = 0.5, fill =
"yellow") +
      geom_area(data = subset(df, x >= upper_bound), alpha = 0.5, fill = "red") +
      labs(x = title, y = "Posterior density of μ") +
      theme_minimal()
  } else {
    ggplot(df, aes(x, y)) +
      geom_area(data = subset(df, x <= lower_bound), alpha = 0.5, fill = "red") +
      geom_area(data = subset(df, x >  lower_bound & x < upper_bound), alpha = 0.5, fill =
"yellow") +
      geom_area(data = subset(df, x >= upper_bound), alpha = 0.5, fill = "green") +
      labs(x = title, y = "Posterior density of μ") +
      theme_minimal()
  }
}
```

```
# uw-mRS mean difference
uw_es <- sapply(seq_along(intervention_counts), function(i) {
  sum(intervention_counts[[i]] * weights) / sum(intervention_counts[[i]]) -
    sum(control_counts[[i]]      * weights) / sum(control_counts[[i]])
})

uw_se <- sqrt(sapply(seq_along(intervention_counts), function(i) {
  var(rep(weights, intervention_counts[[i]])) / sum(intervention_counts[[i]]) +
  var(rep(weights, control_counts[[i]]))      / sum(control_counts[[i]])
}))

uwmrs_df <- data.frame(study = study_labels, es = uw_es, se = uw_se)
cat("uw-mRS (utility-weighted) mean differences:\n"); print(uwmrs_df, row.names = FALSE)

fit_uw <- bayesmeta(
  y = uw_es, sigma = uw_se, labels = study_labels,
  mu.prior.mean = 0, mu.prior.sd = 1
)
cat("\nuw-mRS bayesmeta summary:\n"); print(summary(fit_uw))

# Posterior probabilities for thresholds
p_uw_le_n09 <- fit_uw$pposterior(mu = -0.09)
p_uw_mid    <- fit_uw$pposterior(mu =  0.09) - fit_uw$pposterior(mu = -0.09)
p_uw_ge_p09 <- 1 - fit_uw$pposterior(mu = 0.09)
p_uw_gt_0   <- 1 - fit_uw$pposterior(mu = 0)
cat("\nPosterior probabilities (uw-mRS):\n")
cat("P(mu <= -0.09) =", p_uw_le_n09, "\n")
cat("P(-0.09 < mu < 0.09) =", p_uw_mid, "\n")
cat("P(mu >= 0.09) =", p_uw_ge_p09, "\n")
cat("P(mu > 0) =", p_uw_gt_0, "\n")

plot_posterior_areas_bayesmeta(
  fit_uw, lower_bound = -0.09, upper_bound =  0.09,
  title = "uw-mRS Mean Difference (Thrombolysis – Control)"
```

```
)

# Absolute Risk Difference (ARD) for “minor disability” (mRS 0–1)
ard_es <- sapply(seq_along(intervention_counts), function(i) {
  n_t <- sum(intervention_counts[[i]])
  n_c <- sum(control_counts[[i]])
  pt  <- sum(intervention_counts[[i]][1:2]) / n_t
  pc  <- sum(control_counts[[i]][1:2])      / n_c
  pt - pc
})

ard_se <- sqrt(sapply(seq_along(intervention_counts), function(i) {
  n_t <- sum(intervention_counts[[i]])
  n_c <- sum(control_counts[[i]])
  pt  <- sum(intervention_counts[[i]][1:2]) / n_t
  pc  <- sum(control_counts[[i]][1:2])      / n_c
  pt*(1-pt)/n_t + pc*(1-pc)/n_c
}))

ard_df <- data.frame(study = study_labels, es = ard_es, se = ard_se)
cat("\nAbsolute Risk Difference (mRS 0–1):\n"); print(ard_df, row.names = FALSE)

fit_ard <- bayesmeta(
  y = ard_es, sigma = ard_se, labels = study_labels,
  mu.prior.mean = 0, mu.prior.sd = 1
)
cat("\nARD bayesmeta summary:\n"); print(summary(fit_ard))

p_ard_le_n03 <- fit_ard$pposterior(mu = -0.03)
p_ard_mid    <- fit_ard$pposterior(mu =  0.03) - fit_ard$pposterior(mu = -0.03)
p_ard_ge_p03 <- 1 - fit_ard$pposterior(mu = 0.03)
p_ard_gt_0   <- 1 - fit_ard$pposterior(mu = 0)
cat("\nPosterior probabilities (ARD):\n")
cat("P(mu <= -0.03) =", p_ard_le_n03, "\n")
```

```
cat("P(-0.03 < mu < 0.03) =", p_ard_mid, "\n")
cat("P(mu >= 0.03) =", p_ard_ge_p03, "\n")
cat("P(mu > 0) =", p_ard_gt_0, "\n")

plot_posterior_areas_bayesmeta(
  fit_ard, lower_bound = -0.03, upper_bound =  0.03,
  title = " Minor Disability Absolute Risk Difference (Thrombolysis – Control)"
)

# Mortality Risk Difference (mRS 6)
mort_es <- sapply(seq_along(intervention_counts), function(i) {
  n_t <- sum(intervention_counts[[i]])
  n_c <- sum(control_counts[[i]])
  pt  <- tail(intervention_counts[[i]], 1) / n_t
  pc  <- tail(control_counts[[i]], 1)      / n_c
  as.numeric(pt - pc)
})

mort_se <- sqrt(sapply(seq_along(intervention_counts), function(i) {
  n_t <- sum(intervention_counts[[i]])
  n_c <- sum(control_counts[[i]])
  pt  <- tail(intervention_counts[[i]], 1) / n_t
  pc  <- tail(control_counts[[i]], 1)      / n_c
  as.numeric(pt*(1-pt)/n_t + pc*(1-pc)/n_c)
}))

mort_df <- data.frame(study = study_labels, es = mort_es, se = mort_se)
cat("\nMortality Risk Difference (mRS 6):\n"); print(mort_df, row.names = FALSE)

fit_mort <- bayesmeta(
  y = mort_es, sigma = mort_se, labels = study_labels,
  mu.prior.mean = 0, mu.prior.sd = 1
)
cat("\nMortality bayesmeta summary:\n"); print(summary(fit_mort))
```

```r
p_mort_le_n01 <- fit_mort$pposterior(mu = -0.01)
p_mort_mid    <- fit_mort$pposterior(mu =  0.01) - fit_mort$pposterior(mu = -0.01)
p_mort_ge_p01 <- 1 - fit_mort$pposterior(mu = 0.01)
p_mort_gt_0   <- 1 - fit_mort$pposterior(mu = 0)
cat("\nPosterior probabilities (Mortality RD):\n")
cat("P(mu <= -0.01) =", p_mort_le_n01, "\n")
cat("P(-0.01 < mu < 0.01) =", p_mort_mid, "\n")
cat("P(mu >= 0.01) =", p_mort_ge_p01, "\n")
cat("P(mu > 0) =", p_mort_gt_0, "\n")

plot_posterior_areas_bayesmeta(
  fit_mort, lower_bound = -0.01, upper_bound =  0.01,
  title = "Mortality Risk Difference (Thrombolysis – Control)",
  reverse_colors = TRUE
)

# Forest plots
forestplot(fit_uw,
        main = "uw-mRS Mean Difference (Thrombolysis – Control)",
        xlog = FALSE, predict = TRUE, shrink = FALSE, digits = 2)

forestplot(fit_ard,
        main = "ARD for Minor Disability (mRS 0–1) (Thrombolysis – Control)",
        xlog = FALSE, predict = TRUE, shrink = FALSE, digits = 2)

forestplot(fit_mort,
        main = "Mortality Risk Difference (Thrombolysis – Control)",
        xlog = FALSE, predict = TRUE, shrink = FALSE, digits = 2)
```

**APPENDIX 3: Forest plots**

| study | estimate | 95% CI |
|---|---|---|
| ECASS-4 (Alteplase) | 0.070 | [-0.100, 0.240] |
| EXTEND (Alteplase) | 0.059 | [-0.063, 0.181] |
| THAWS (Alteplase) | -0.012 | [-0.187, 0.163] |
| WAKE-UP (Alteplase) | 0.113 | [0.025, 0.201] |
| TRACE-3 (Tenecteplase) | 0.087 | [0.010, 0.165] |
| HOPE (Alteplase) | 0.140 | [0.045, 0.235] |
| **mean** | **0.091** | **[0.026, 0.152]** |
| **prediction** | **0.091** | **[-0.041, 0.215]** |

*Heterogeneity (tau): 0.029 [0.000, 0.111]*

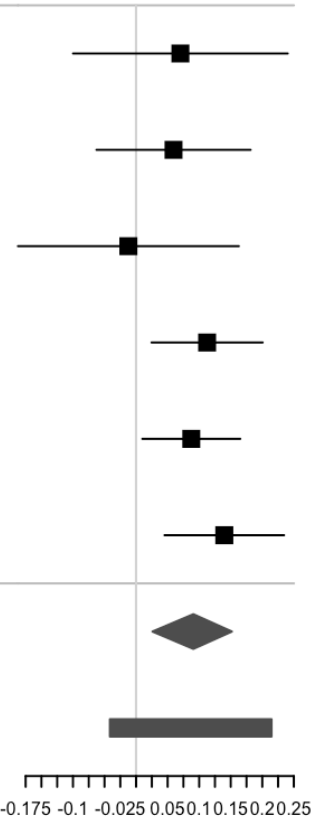


SFigure 4. Bayesian random-effects meta-analysis for a minor disability outcome (Thrombolytic – Control).

| study | estimate | 95% CI |
|---|---|---|
| ECASS-4 (Alteplase) | 0.047 | [-0.059, 0.154] |
| EXTEND (Alteplase) | 0.026 | [-0.053, 0.105] |
| THAWS (Alteplase) | -0.005 | [-0.067, 0.057] |
| WAKE-UP (Alteplase) | 0.033 | [0.005, 0.060] |
| TRACE-3 (Tenecteplase) | 0.002 | [-0.057, 0.060] |
| HOPE (Alteplase) | 0.000 | [-0.063, 0.063] |
| **mean** | **0.019** | **[-0.016, 0.050]** |
| **prediction** | **0.019** | **[-0.050, 0.083]** |

*Heterogeneity (tau): 0.016 [0.000, 0.057]*

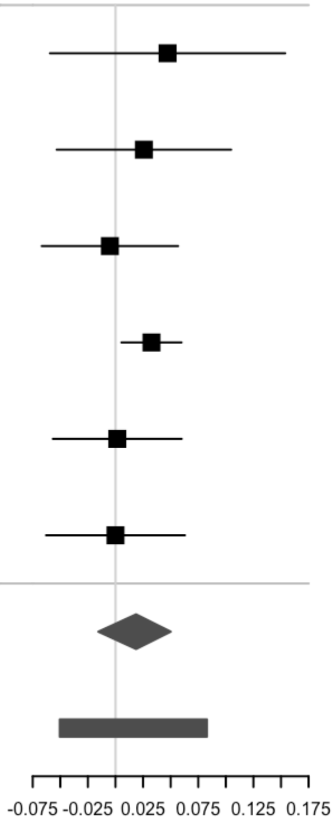


SFigure 5. Bayesian random-effects meta-analysis for a mortality outcome (Thrombolytic – Control)

**APPENDIX 4: Sensitivity analysis**

4.1 The posterior effect size applying a weakly informative prior effect size ($\mu$=0, sd=.04) was .04 (95% CrI, 0.0 - .07). The posterior probability of any benefit was .97; clinically significant benefit was~0; clinical indifference was ~.99; and clinical harm was ~0.

4.2 The posterior effect sizes utilizing different utility weights were similar (STable 1).

| Weights | Estimate | 95% Credible Interval |
|---|---|---|
| DAWN | .05 | .01 - .08 |
| BEST-MSU | .05 | .01 - .09 |
| Wang et al. | .05 | .01 - .09 |

STable 2. Sensitivity analysis using weights from the BEST-MSU study and study by Wang et al. [2,3]

| *Author (Year)* | *PICO* | *Prespecified Protocol* | *Performed Comprehensive Search* | *Assessed Risk of Bias* | *Appropriate Meta-analytical Method* | *ROB Consideration in Interpretation* | *Publication Bias Assessment* |
|---|---|---|---|---|---|---|---|
| Tsivgoulis (2020) | Patients: Acute Ischemic Stroke beyond 4.5 hrs; Interventions: Thrombolysis (alteplase only) with penumbral and DWI/FLAIR mismatch; Comparator: Placebo or standard care; Outcomes: mRS 0–1, mRS 0–2 | Not referenced in main text | Yes | Yes | Yes | No | Yes |
| Campbell (2019) | Patients: Acute Ischemic Stroke between 4.5–9 hrs and upon awakening; Interventions: Thrombolysis (alteplase only) with penumbral mismatch; Comparator: Placebo or standard care; Outcomes: mRS 0–1, mRS 0–2, mRS 0–6 | Yes | Yes | Yes | Yes | No | Yes |
| Mac Grory (2021) | Patients: Acute Ischemic Stroke upon awakening; Interventions: Thrombolysis (alteplase only) with DWI/FLAIR mismatch; Comparator: Placebo or standard care; Outcomes: mRS 0–1, mRS 0–2 | Yes | Yes | Yes | Yes | No | Yes |
| Thomalla (2020) | Patients: Acute Ischemic Stroke with unknown onset; Interventions: Thrombolysis (alteplase only) with penumbral or DWI/FLAIR mismatch; | Yes | Yes | Yes | Yes | No | Yes |

| | | | | | | | |
|---|---|---|---|---|---|---|---|
| | Comparator: Placebo or standard care; Outcomes: mRS 0–1, mRS 0–2, mRS 0–3, mRS 4–6, mRS 0–6 | | | | | | |
| Gunkan (2025) | Patients: Acute Ischemic Stroke beyond 4.5 hrs; Interventions: Thrombolysis (alteplase or tenecteplase) with penumbral or DWI/FLAIR mismatch; Comparator: Placebo or standard care; Outcomes: mRS 0–1, mRS 0–2, mRS 0–6 (graphical only) | Yes | Yes | Yes | Yes | No | Yes |
| Roaldsen (2021) | Patients: Acute Ischemic Stroke upon awakening; Interventions: Thrombolysis (alteplase only) with penumbral or DWI/FLAIR mismatch; Comparator: Placebo or standard care with penumbral or DWI/FLAIR mismatch; Outcomes: mRS 0–2 | Yes | Yes | Yes | Yes | No | Yes |
| Jia (2021) | Patients: Acute Ischemic Stroke beyond 4.5 hrs; Interventions: Thrombolysis (alteplase only) with penumbral or DWI/FLAIR mismatch; Comparator: Placebo or standard care; Outcomes: mRS 0–1, mRS 0–2 | Not referenced in main text | No | Yes | Yes | No | Yes |

STable 3. AMSTAR 2 Critical Reporting Domains for included meta-analyses.

| *Author (Year)* | *Critical Outcome* | *Assessed Risk of Bias* | *Risk of Bias Due to Early Termination* | *Risk of Bias Due to Financial Conflicts of Interest* | *Assessed Inconsistency* | *Assessed Imprecision* | *MCID Defined* | *Assessed Publication Bias* | *Inclusion of Phase-2 RCTs < 100 Participants Per Group* | *Subgroups of Participants Included* |
|---|---|---|---|---|---|---|---|---|---|---|
| Tsivgoulis (2020) | mRS 0-1 | Yes | Unclear | Not Acknowledged | Yes | No | No | Yes | Yes | No |
| Campbell (2019) | mRS 0-1 | Yes | Unclear | Not Acknowledged | Yes | No | No | Yes | Yes | Yes |
| Mac Grory (2021) | mRS 0-2 | Yes | Not Acknowledged | Not Acknowledged | Yes | No | No | Yes | No | No |

| | | | | | | | | | | |
|---|---|---|---|---|---|---|---|---|---|---|
| Thomalla (2020) | mRS 0-1 | Yes | Not Acknowledged | Not Acknowledged | Yes | No | No | Yes | No | Yes |
| Gunkan (2025) | mRS 0-1 | Yes | Not Acknowledged | Not Acknowledged | Yes | No | No | Yes | Yes | No |
| Roaldsen (2021) | mRS 0-2 | Yes | Unclear | Not Acknowledged | Yes | No | No | Yes | Yes | Yes |
| Jia (2021) | mRS 0-1 | Yes | Not Acknowledged | Not Acknowledged | Yes | No | No | Yes | Yes | No |

STable 4. GRADE domains for included meta-analyses.
